# Monitoring and forecasting the COVID-19 epidemic in Moscow: model selection by balanced identification technology - version: September 2021

**DOI:** 10.1101/2021.10.07.21264713

**Authors:** Alexander Sokolov, Lyubov Sokolova

## Abstract

A mathematical model is a reflection of knowledge on the real object studied. The paper shows how the accumulation of data (statistical data and knowledge) about the COVID-19 pandemic lead to gradual refinement of mathematical models, to the expansion of the scope of their use. The resulting model satisfactorily describes the dynamics of COVID-19 in Moscow from 19.03.2020 to 01.09.2021 and can be used for forecasting with a horizon of several months. The dynamics of the model is mainly determined by herd immunity. Monitoring the situation in Moscow has not yet (as of 01.09.2021) revealed noticeable seasonality of the disease nor an increase in infectivity (due to the Delta strain). The results of using balanced identification technology to monitor the COVID-19 pandemic are:

- models corresponding to the data available at different points in time (from March 2020 to August 2021);
- new knowledge (dependencies) acquired;
- forecasts for the third and fourth waves in Moscow.

Discrepancies that manifested after 01.09.2021 and possible further modifications of the model are discussed

## Introduction

Effective monitoring and forecasting of the behaviour of complex objects implies the extensive use of mathematical modelling. The availability of mathematical models should be factored in when determining the goals of monitoring and forecasting. The complexity, detail and reliability of said models, in turn, is determined by the amount and quality (accuracy) of experimental data and knowledge about the functioning of the object.

In case of the COVID-19 pandemic, as data and knowledge accumulated, and more detailed and accurate models were built, monitoring and forecasting capabilities increased. Various effects appeared in the available data series, which allowed to complicate the corresponding mathematical models. This complication was also facilitated by new knowledge about the processes under study.

The dynamics of an epidemic is determined by the processes of interaction between the virus, the human body and the society. Different processes have different characteristic times, and the longer the characteristic time of a process, the longer it takes for its manifestation, the longer are the series of observations necessary to determine its characteristics.

For the COVID-19 pandemic characteristic times may be estimated as:

- 15 days – infectiousness and manifestation (detectability) as a function of the duration of the disease;
- 30 days – current reproductive index (contact number) and index of detection (and subsequent isolation);
- 60 days –the detection index as a function of the number of tests performed;
- 90 days – the influence of natural (after illness) herd immunity;
- 200 days – the effect of natural immunity weakening over time;
- 100 days – impact of vaccination;
- 180 days (preliminary estimate) – the effect of artificial (vaccination caused) immunity weakening;
- 365 days – seasonal infectivity.

The forecast horizon is determined by the model error, which is determined by the accuracy of the individual processes’ description, which in turn is determined by observation series’ length and reliability. Therefore, monitoring timeframe determines the processes that can be estimated (with characteristic times below said timeframe), thus defining forecast horizon and precision. This may explain the failure of N. Ferguson’s efforts to predict the dynamics of the pandemic. In the beginning of 2020 his group was keenly justifying complete quarantine, predicting (with a horizon of a few months) no less than 500 thousand deaths in Great Britain and 2 million in the United States (by the end of summer 2020). [https://www.imperial.ac.uk/media/imperial-college/medicine/mrc-gida/2020-03-16-COVID19-Report-9.pdf, p. 7]

First few months of the pandemic, when the volume of knowledge and statistical data was small and unreliable, we limited our research to looking for patterns of interaction between the virus and humans (infectivity and manifestation of the virus as a function of the duration of the disease) and describing some of the social mechanisms of epidemic management (the number of contacts and isolation of the infected). Processes with characteristic times of about 30 days were considered.

Then, as experience (analysis of model trajectories and their comparison with the initial data), knowledge and statistical information were accumulated, a number of modifications were carried out, which made it possible to factor in slower processes (with longer characteristic times):

- the effectiveness of isolating the infected as a function of the number of the tests performed (daily) (summer 2020);
- natural immunity of the recovered (autumn 2020);
- loss of immunity as a function of the age of infection (winter 2020-2021);
- vaccination (spring-summer 2021).

## Data sources

Following statistical data was used:

- New cases of infection in Moscow – from official records (see ex., https://стопкоронавирус.рф/information/).
- The number of tests conducted in Russia – from the official reporting (see ex., https://tatspravka.ru/statistika-russia/). Number of tests performed per 1000 people in Moscow was estimated at 1/5 of those held in Russia.

- Percent of immunized people (with active detectable antibodies) in Moscow – from government officials’ statements and internet citations:
- https://ria.ru/20200710/1574177822.html,
- https://www.vedomosti.ru/society/news/2020/09/28/841417-popova-nazvala-dolyu-moskvicheis-antitelami-k-koronavirusu,

- https://www.vesti.ru/article/2533510
- The number of vaccinations carried out in Moscow – from the official records, (see ex., https://стопкоронавирус.рф/news/)

## Methods and models

The method of balanced identification [Sokolov 2020a] and the corresponding information technology of the same name, created at the Center for Distributed Computing of the Institute for Information Transmission Problems of the Russian Academy of Sciences, were used to select models corresponding to the quantity and quality of data. A software implementation of the technology is currently available (https://github.com/distcomp/SvF).

It should be noted that the main variable of the model, the number of undetected infected, is not an observable (measurable) value. The observed value is newly detected cases of infection (*nC*), which are an unknown function of the undetected. This leads to the ambiguity of identification as the internal dynamics of the model is determined up to a factor. To overcome this shortcoming, a special term minimizing the total number of infected is added to the criterion for balanced identification. In addition, to restore the correct problem statement (elimination of the ambiguity of identification), supplementary data is used – the number of people with antibodies (obtained by integrating the total number of infected with immunity).

Various modifications of the *SIR* and *SIRS* models are used as the accepted designations of epidemiological models (see e.g., [Brauer 2019]), with a prominent feature of dividing the infected (*I*) into 15 groups (Age of Infection Model, see e.g. *SI*_*15*_*R* in the Appendix) by age of infection (the duration of the disease). In terms of population dynamics and demography models, Leslie’s (matrix) models [Svirezhev 1978] and the McKendrick–Von Foerster’s models for populations with an age structure [Nakhushev 1995, Ebeling 2001] are used.

The structure of the models changed as new data became available and new processes were factored in (testing, vaccination, etc.). The specific “character-numeric” designations of the respective model revisions reflect the data used and the processes accounted for. Formal descriptions of the models are given in the Appendix.

## Results

At the beginning of the pandemic we knew almost nothing about the virus, and the available statistical information was limited to the numbers of new cases of infection (*nC*) and deaths. At this stage, we limited ourselves to identifying the patterns of virus-human interaction – infectiousness and manifestation as functions of the duration of the disease. For this, we used a model of the dynamics of undetected infected *SI*_*15*_*R-nC* (see Appendix), distributed over the duration of infection, built using only the time series of detected infected (*nC*). After a couple of months, based on time series for 7 populations (Great Britain, Germany, Italy, Spain, France, Russia without the city of Moscow and the Moscow region and the city of Moscow with the Moscow region), we assessed the functions of human-virus interaction that determine the dynamics of the epidemic, for example, infectiousness as a function of the duration (time) of the disease (see Fig. 1), and social controls – the current reproductive index *R0*_*t*_ (contact number) and the index of detection (and subsequent isolation) *IS*_*t*_ (For further information, see [Sokolov 2020]).

**Fig. 1.**
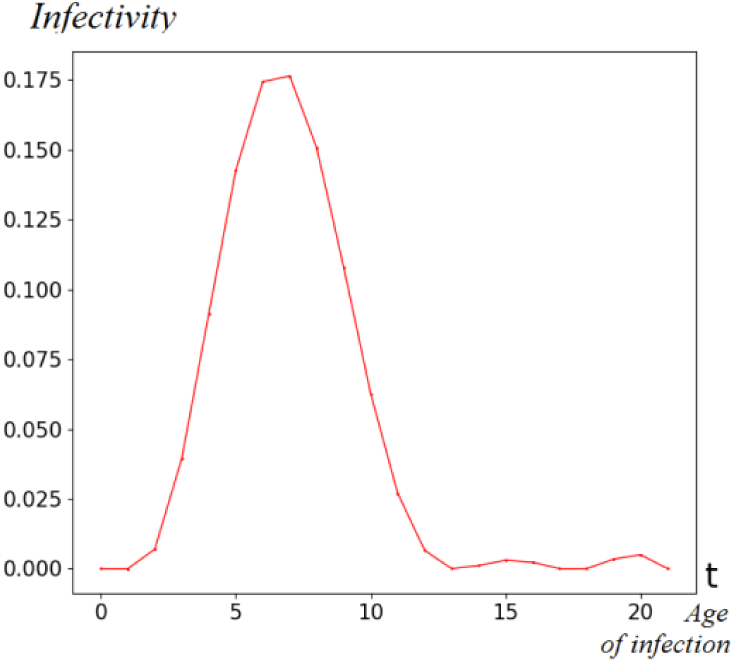
Normalized (integral equals one) infectivity as a function age of infection (days).

By summer 2020, a sufficient amount of new information – the number of tests performed – has been accumulated. This allowed us to set up a new goal – to describe the efforts of society to identify (and subsequently isolate) patients as a function of the number of tests performed. The corresponding modified model *SI*_*15*_*R-nC-nT* (see Appendix) made it possible to assess the relationship between the number of detected infected and the number of tests performed. Taking into account asymptomatic and unregistered carriers of the virus, the detection efficiency as a function of the number of tests per thousand people for Moscow is shown in Fig. 2. Analysis of the curve allows to evaluate the efficiency of detecting patients in Moscow for this model as no more than 17%, i.e. of the total number of infected, no more than one in six is identified and isolated.

**Fig. 2.**
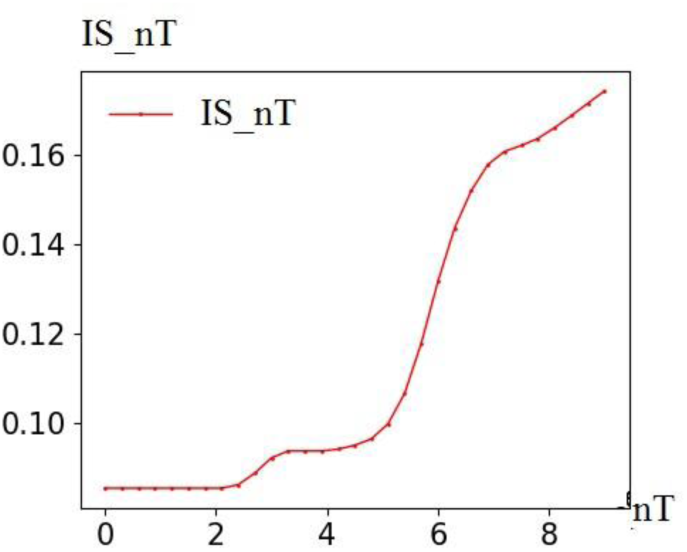
*IS_nT* – a function of the effectiveness of detecting the infected depending on the number of tests per thousand (*nT*) in Moscow.

This modification can already be used to predict morbidity under various scenarios of volume of testing.

These two models were linear, they determined the internal dynamics up to a factor and did not take into account the population size. The real number of infected is unknown (not an observable value). A new type of information was needed to estimate the total number of infected.

In the summer of 2020 this information appeared – (screening) estimates of the number of carriers of antibodies (with significant antibody titre), acquired by taking integral of the total number of infected (not only detected infected). This (non-systematic) information was obtained from the government officials’ statements for the population of Moscow. The corresponding modification of the model – *SI*_*15*_*R-nC-nT-Anti* – allowed to estimate the real number of infected (not only the detected cases) in Moscow and to factor in the number of those who had recovered (they are believed to have natural immunity), which significantly improved the predictive capabilities of the model. The model was no longer linear, self-regulation of the process appeared.

However, in December 2020 the solution space of the model became insufficient for the observed data, which caused its next modification – factoring in the loss of natural (i.e. obtained as a result of the disease) immunity. The identification of a new modification of the *SI*_*15*_*RS-nC-nT-Anti-Im* model allowed to estimate the time of immunity loss at 190 days (on average) – the corresponding curve is shown in Fig. 3.

**Fig. 3.**
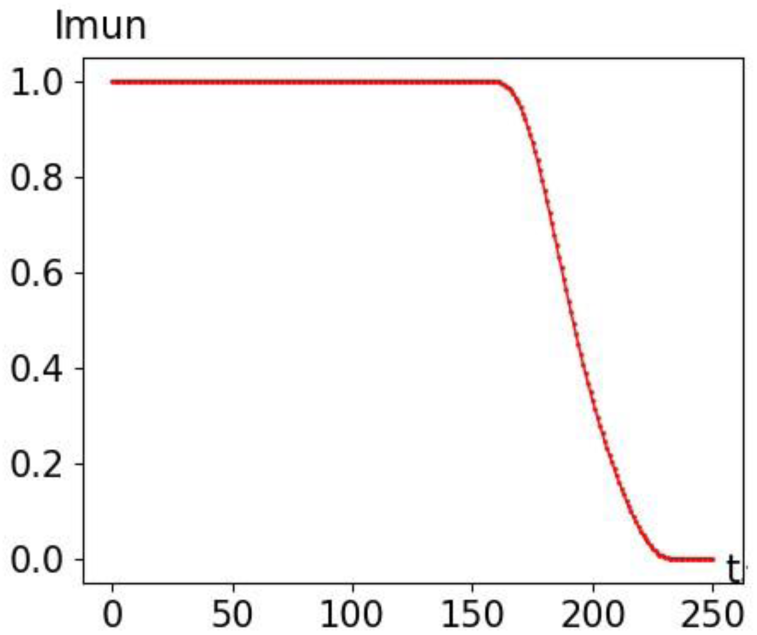
Preservation of natural immunity as a function of time from the onset of the disease (days). 1 – total immunity, 0 – no immunity.

Finally, at the beginning of 2021, mass vaccination began. To factor it in the modification of the *SI15RS-nC-nT-Anti-Im-Vuc* model we added a multiplier that reduces the number of new infections depending on the percentage of vaccinated. In this modification, there are already three (social) controls for epidemic management: vaccination, testing and contact restriction.

### Forecasts

By December 2020, sufficient monitoring experience has been accumulated, and the main processes describing the dynamics of the pandemic have been discovered, described and identified. The resulting models were used for forecasting.

Figures 4, 5, 7, 8 and 12 show the results of modeling, identification, prediction and verification of new (identified) cases. The X-axis represents the number of days from the start of the simulation – 19.03.2020. For clarity, the marks for the months and years are shown above the X-axis. The vertical yellow line corresponds to the forecast start date. Model curves (in red) were obtained (identified) on the training dataset (blue dots). Then the model was used for forecasting. At the same time, the social controls of the model (the replication index, the number of new tests and the number of newly vaccinated) were frozen at the last value before the forecast. To verify the model, the data set obtained later and not used in the construction of the model (green dots) was utilized. Comparison of the forecast (red curve) with the test set shows good agreement over several months. The resulting discrepancies, as a rule, can be explained by significant changes of social controls.

**Fig. 4.**
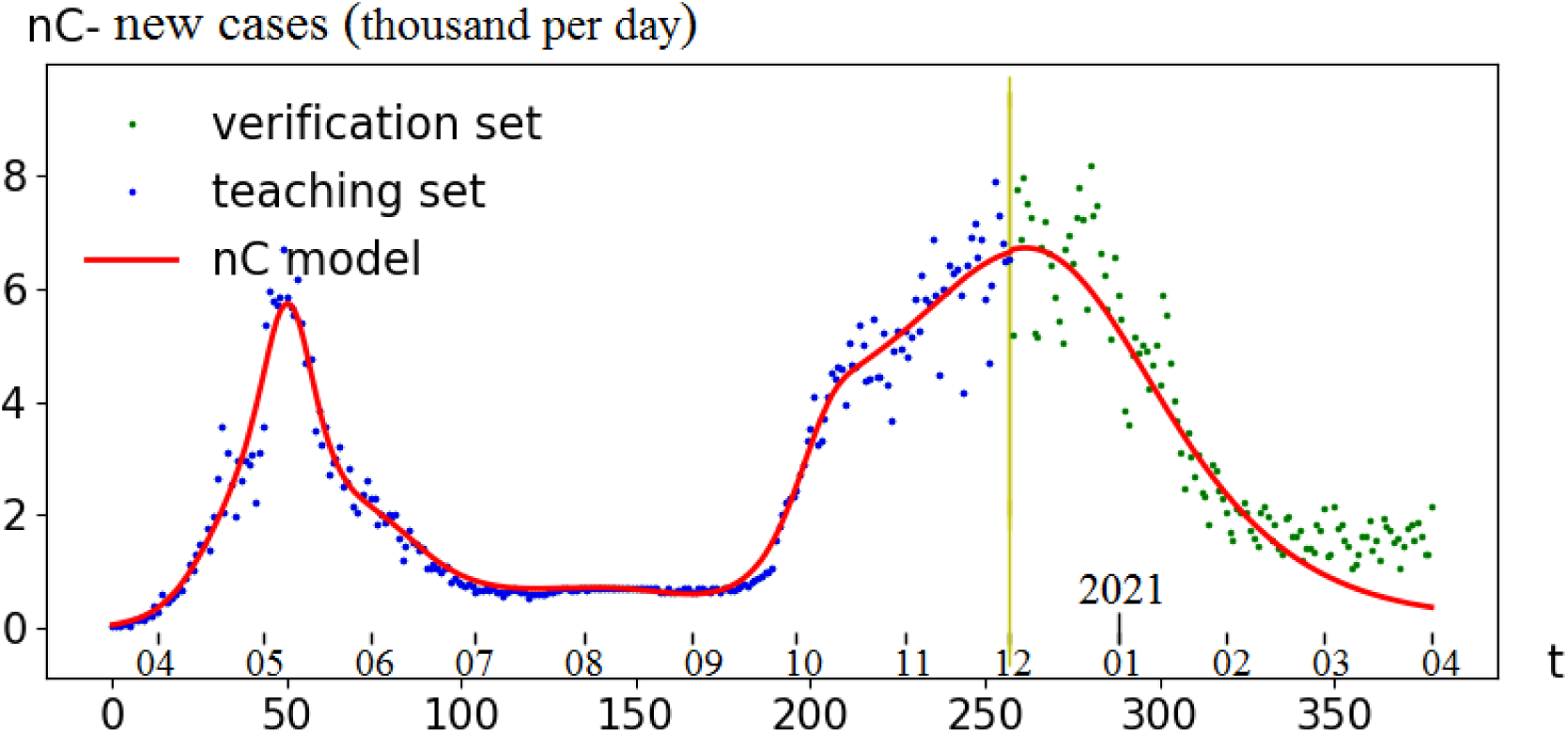
Forecast of new cases of infection in Moscow from 01.12.2020. Blue dots – training set, red curve – the model, green dots – testing set.

#### Forecast from 01.12.2020 – end of the second wave

According to the model, the drop in the number of detected cases in the second wave (December-January 2021) is largely determined by the accumulation of the immune layer (the population with natural immunity). The discrepancy observed since mid-February can be explained by the significant weakening of (anti-epidemic) restrictions and/or by migration from other Russian regions.

#### Forecast from 15.04.2021 – the third wave

Fig. 5. shows an example of the forecast of the third wave of pandemic in Moscow with preset controls: contacts and tests are frozen at 2.43 and 4.48 respectively (as of April 15, 2021), vaccination – 12,000 people per day, vaccination efficiency 91.6% (see Fig. 6).

**Fig. 5.**
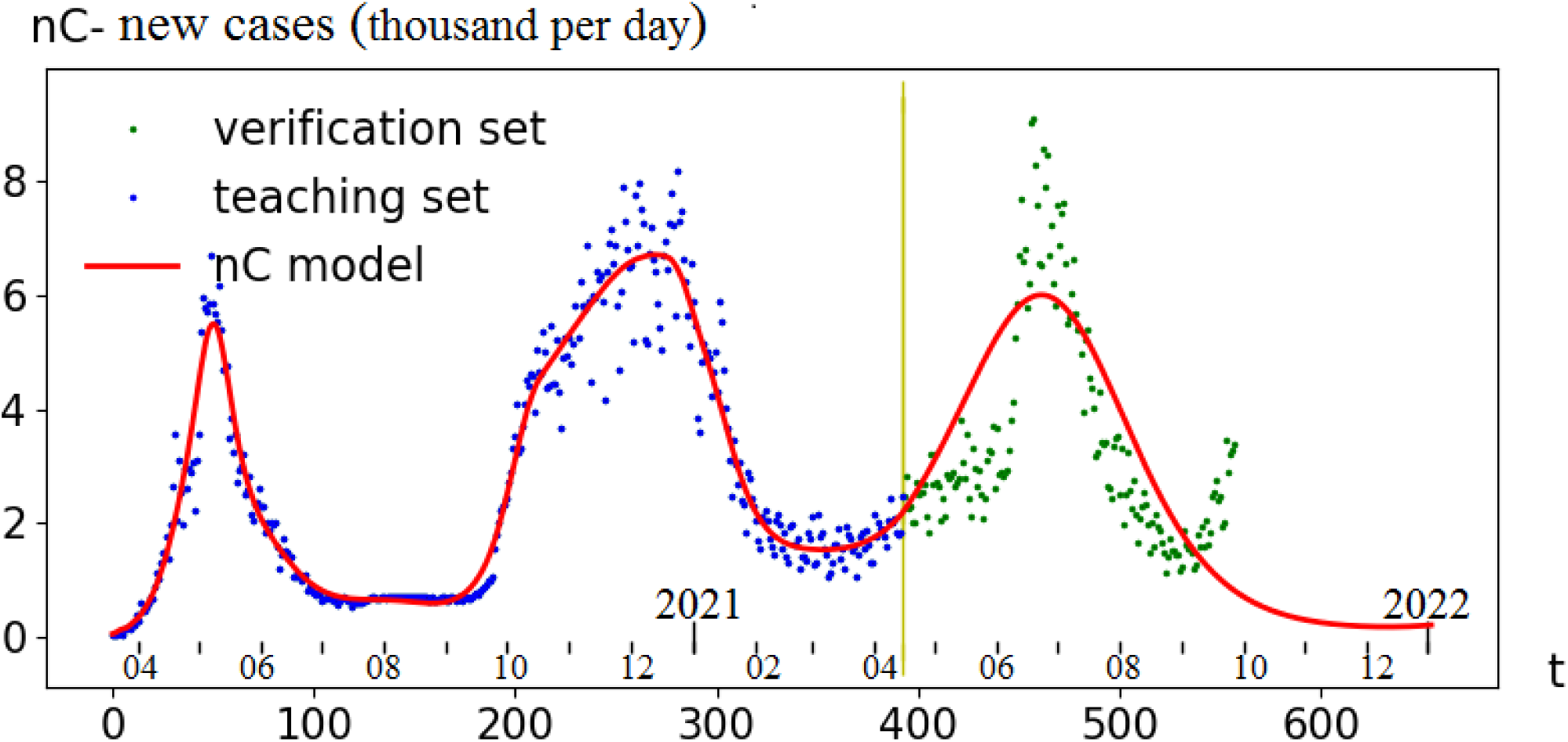
Forecast of the third wave of new cases of infection in Moscow from 15.04.2021. Blue dots – training set, red curve – model, green dots – (verification?) testing set (up until 20.09.2021).

**Fig. 6.**
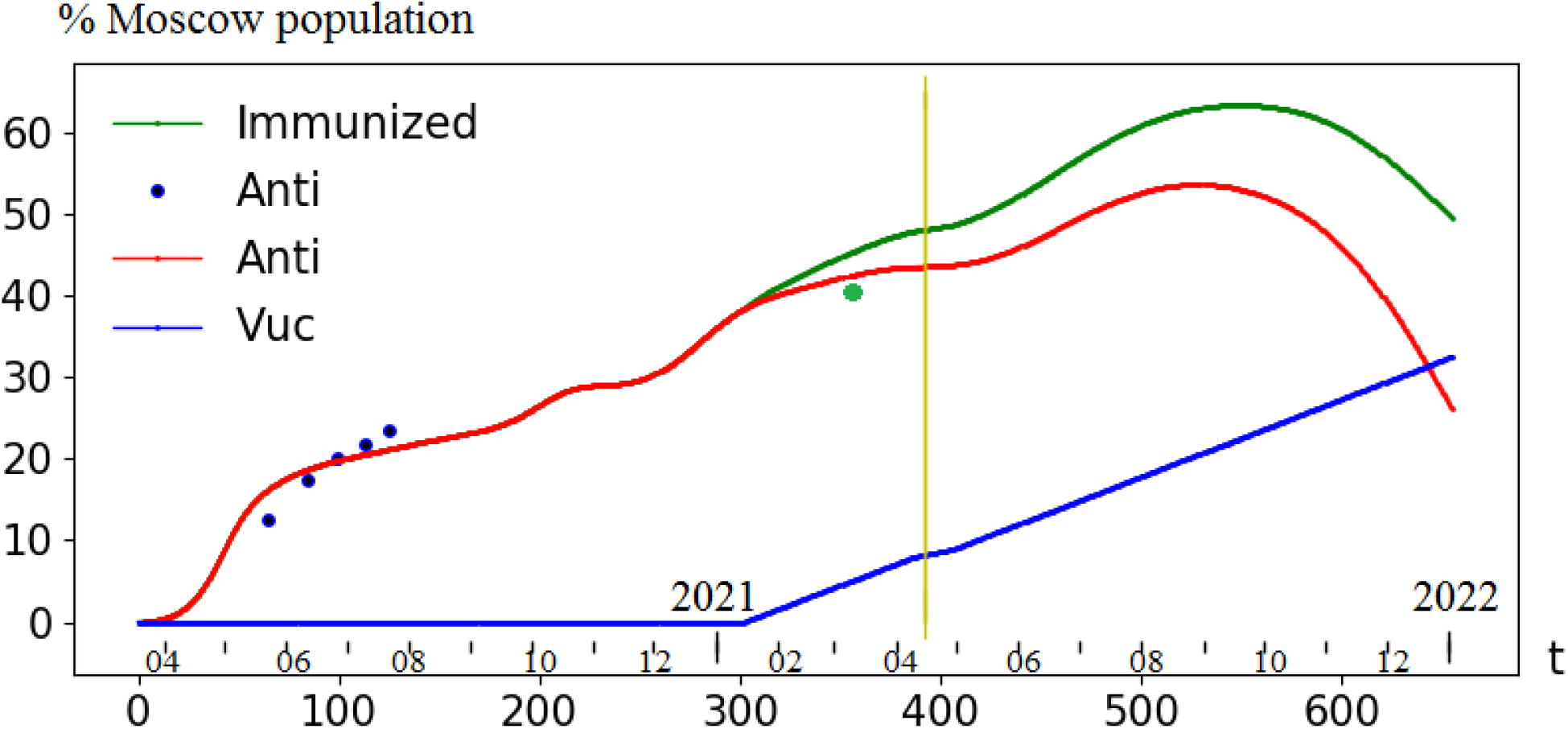
Immunization of the population of Moscow. *Vuc(t)* – preset management, the percentage of vaccinated (corresponds to the rate of vaccination of 12 thousand/day). *Anti(t)* – forecast of the percentage of those recovered who retained their immunity by the time *t. Immunized(t)* – forecast of the percentage of “immunized” (vaccinated and recovered with immunity). Blue dots (percentage of carriers of antibodies, i.e., those who had recovered) were used for identification, the green dot was left for verification.

Over the next five months, the forecast qualitatively coincides with the real data that was not used to identify the model (green dots). According to the model, the third wave (an increase in the number of detected cases from May 2021 on) is largely determined by a decrease in the immune layer of the recovered population. Indeed, since May 2021 those who have been infected at the beginning of the second wave (October-November 2020) started to lose their immunity. In June, the growth of the identified infected increases sharply, which corresponds to the loss of natural immunity by a significant number of those who contracted the virus in December 2020. The waves’ frequency is about 200 days and is mainly determined by the function of preservation of immunity (Fig. 3). Aggressive measures to temporarily decrease the replication index and increase testing can slightly shift the wave or change its shape. But the total number of infected changes insignificantly as the dynamics is mainly determined by the systemic (both of recovered and vaccinated) herd immunity. According to the model (*SI*_*15*_*RS-nCnT-Anti-Im-Vuc*), for the wave to subside, the general herd immunity must exceed a certain value, which is determined by the number of tests and the reproductive index (restrictive measures of society). For the first wave, this value was about 15% of the population, for the second – about 35%, for the third it is estimated at about 55%.

With vaccination rate of 12 thousand per day, the forecast for the number of detected infected in the third wave (from 15.04.2021 to 01.11.2021) is 671 thousand people. The estimated mortality (with a 2.5% mortality rate) is about 16 thousand. The introduction of (temporary) restrictions that reduce *R0*_*t*_ and an increase in testing rates stretch the wave and decrease its height, however, the predicted total number of detected cases in the third wave does not change significantly. This is a “constant” of a kind that can be significantly reduced only by vaccination – according to model calculations (estimations as of June 17, 2021), 11 vaccinated reduce the number of recovered patients by 1. Thus, vaccinating surplus (as compared to the forecast at the rate of 12 thousand per month) 1.5 million people reduces the number of detected cases by 135 thousand. With a mortality rate of 2.5%, 440 vaccinated people reduce death toll in the third wave by 1, thus, 1.5 million additional vaccinated save about 3,400 lives.

According to official statistics, the number of cases detected in Moscow from 15.04.2021 to 19.07.2001 is 410 thousand of people, which is in good agreement with 671 thousand predicted, especially taking into account the 120 thousand decrease due to the intensive vaccination not foreseen by the forecast.

Figure 6. shows the dynamics of herd immunity and its components. The dot with a green circle (estimation of the percentage of patients with immunity as of March 08, 2021) was not used for identification. Its proximity to the trajectory can be considered an additional confirmation of the suitability of the model for forecasting (model verification).

#### Forecast from 11.05.2021

The forecast seen in Fig. 7 (from 11.05.2021) corresponds to the level of contacts typical for holidays (01.05.2021–10.05.2021), which explains the noticeable slowdown in the virus incidence. In this case, the predicted third wave is lower and wider, but has almost the same area (the total number of detected infected) as in the forecast in Fig. 5.

**Fig. 7.**
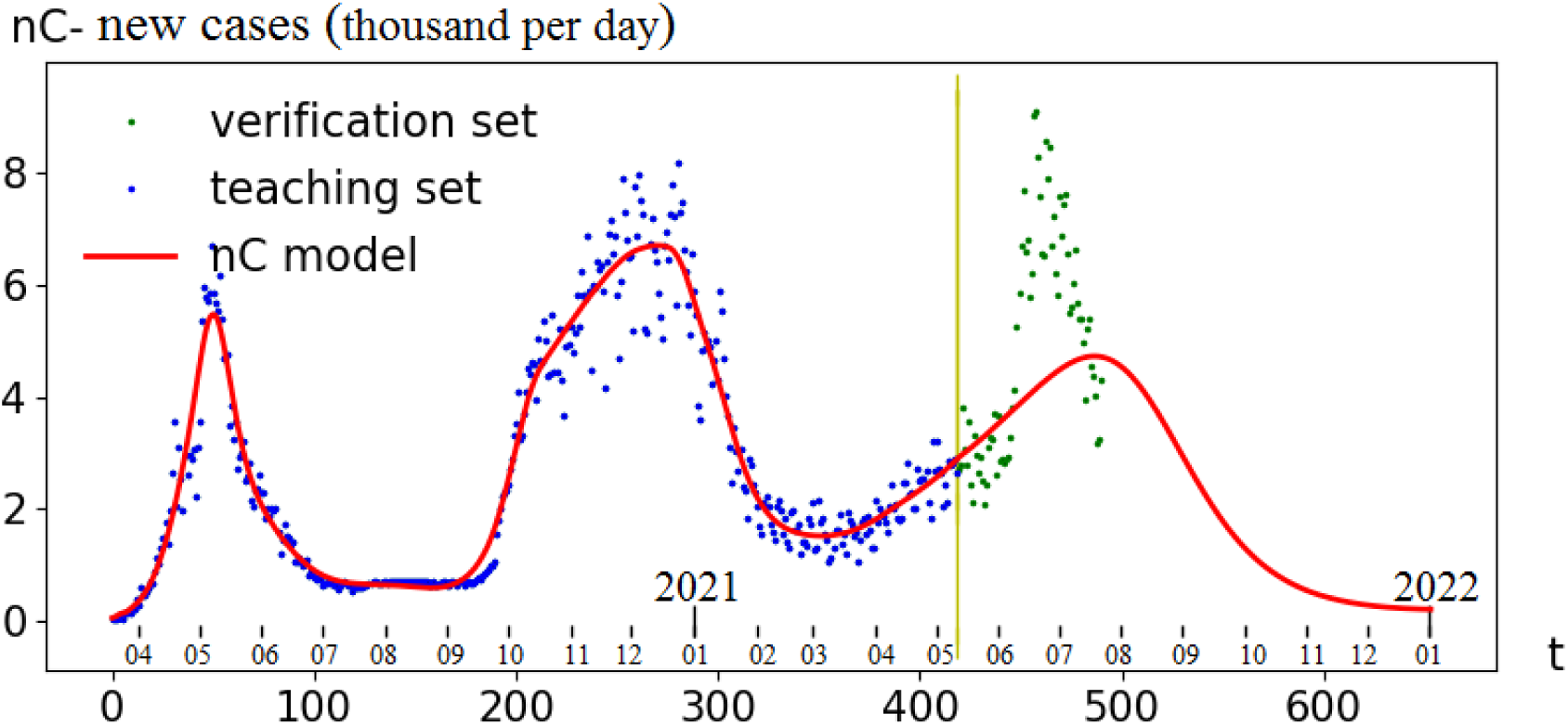
Example of the third wave forecast for Moscow from 11.05.2021.

#### Forecast from 25.07.2021 – end of the third wave

Let’s delve deeper. The calculation of the forecast shown in Fig. 8, was carried out according to the following control scenarios:

**Fig. 8.**
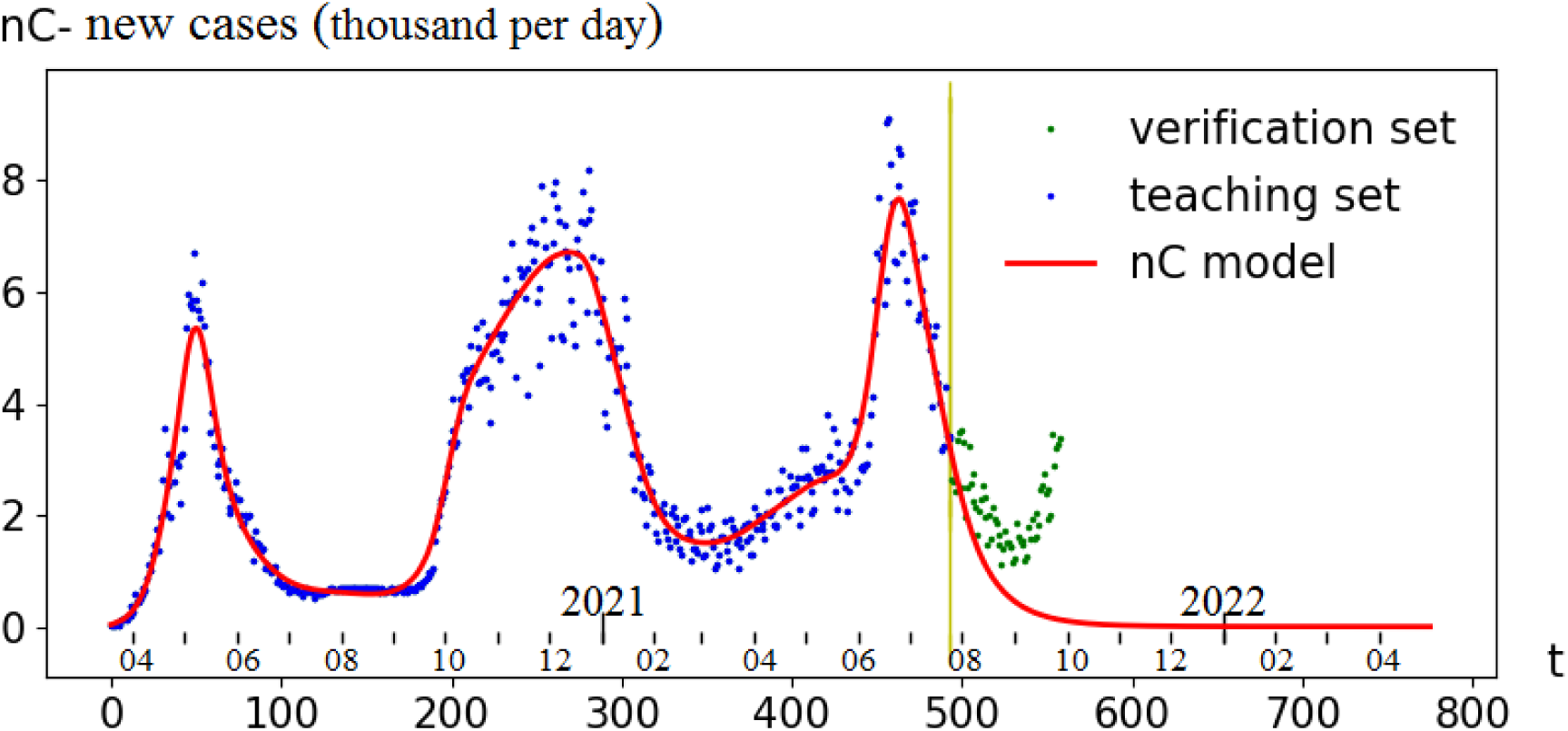
Third wave forecast for Moscow from 25.07.2021.

- Vaccination in % of the population of Moscow (in Fig. 9, the *Vuc* graph, shifted by 2 weeks) based on the intensity of 12 thousand/day, the effectiveness is 91.6,
- the reproduction index (*R0* in Fig. 10) is frozen at the last value *R0* = 1.9,
- the number of tests per thousand (Fig. 11) is frozen at the last value *nT* = 8.7.

**Fig. 9.**
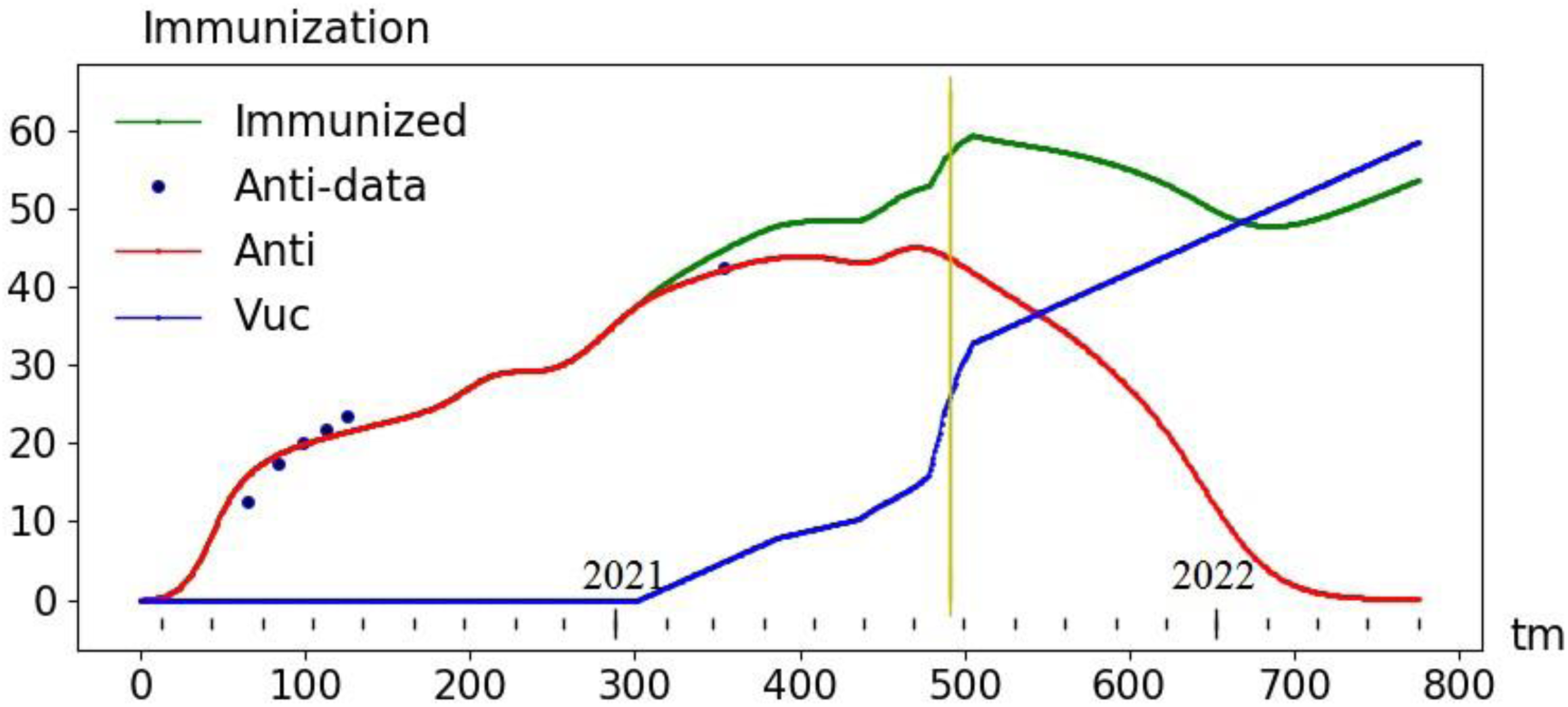
Forecast of immunization of the Moscow population from 25.07.2021. *Vuc(t)* – preset control, the percentage of vaccinated (corresponds to the rate of vaccination of 12 thousand/day). *Anti(t)* – forecast of the percentage of the recovered who retained their immunity by the time *t. Immunized(t)* – forecast of the percentage of “resistant” (vaccinated and recovered with immunity)

**Fig 10.**
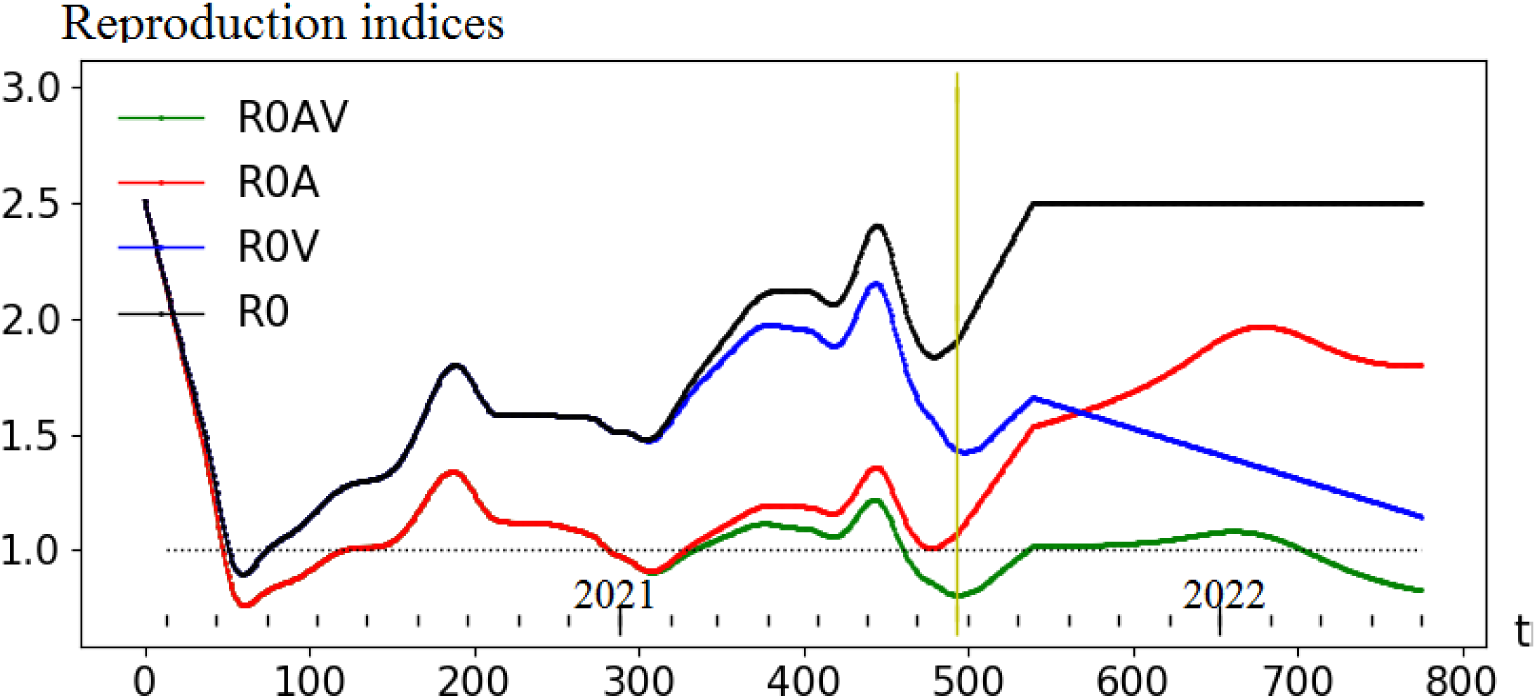
Reproduction indices (as of 25.07.2021): *R0* – reproduction index (identified function and preset control, to the right of the yellow line), *R0A* – reproduction index with the immunity of the recovered factored in, *R0V* – reproduction index with vaccination factored in, *R0AV* – reproduction index with the immunity of the recovered and vaccinated factored in.

**Fig. 11.**
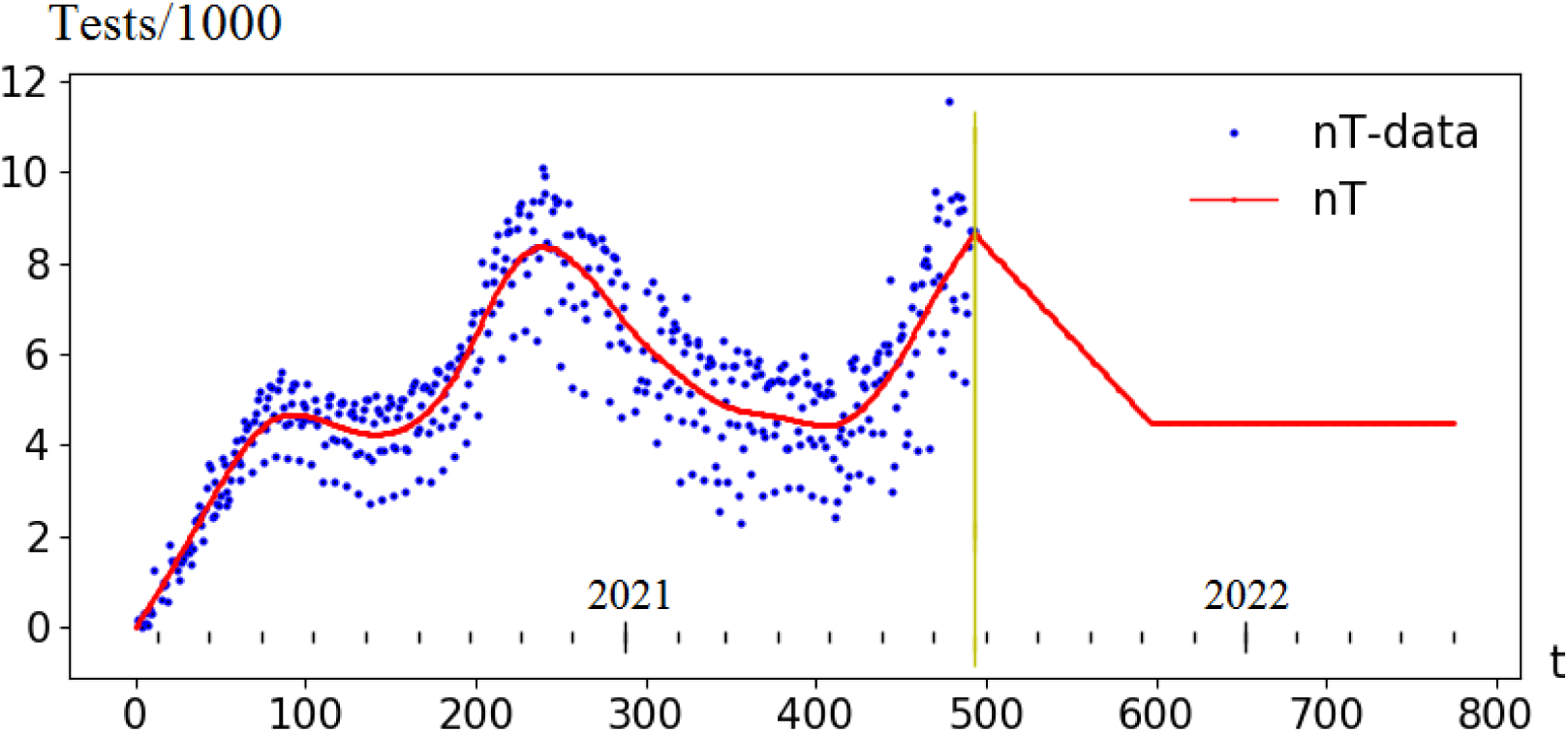
Number of tests per thousand: data fitting and preset control (to the right of the yellow line).

Obviously, the scenarios used (low reproduction index and a significant number of tests) considerably underestimate the forecast results. In this case, even 50% vaccination makes it possible to avoid the fourth wave (by the end of 2021).

#### Forecast from 25.07.2021 – the fourth wave

For more realistic forecast see Fig. 12-15, assuming restrictions alleviation and testing rates reduction while maintaining the same vaccination scenario:

**Fig. 12.**
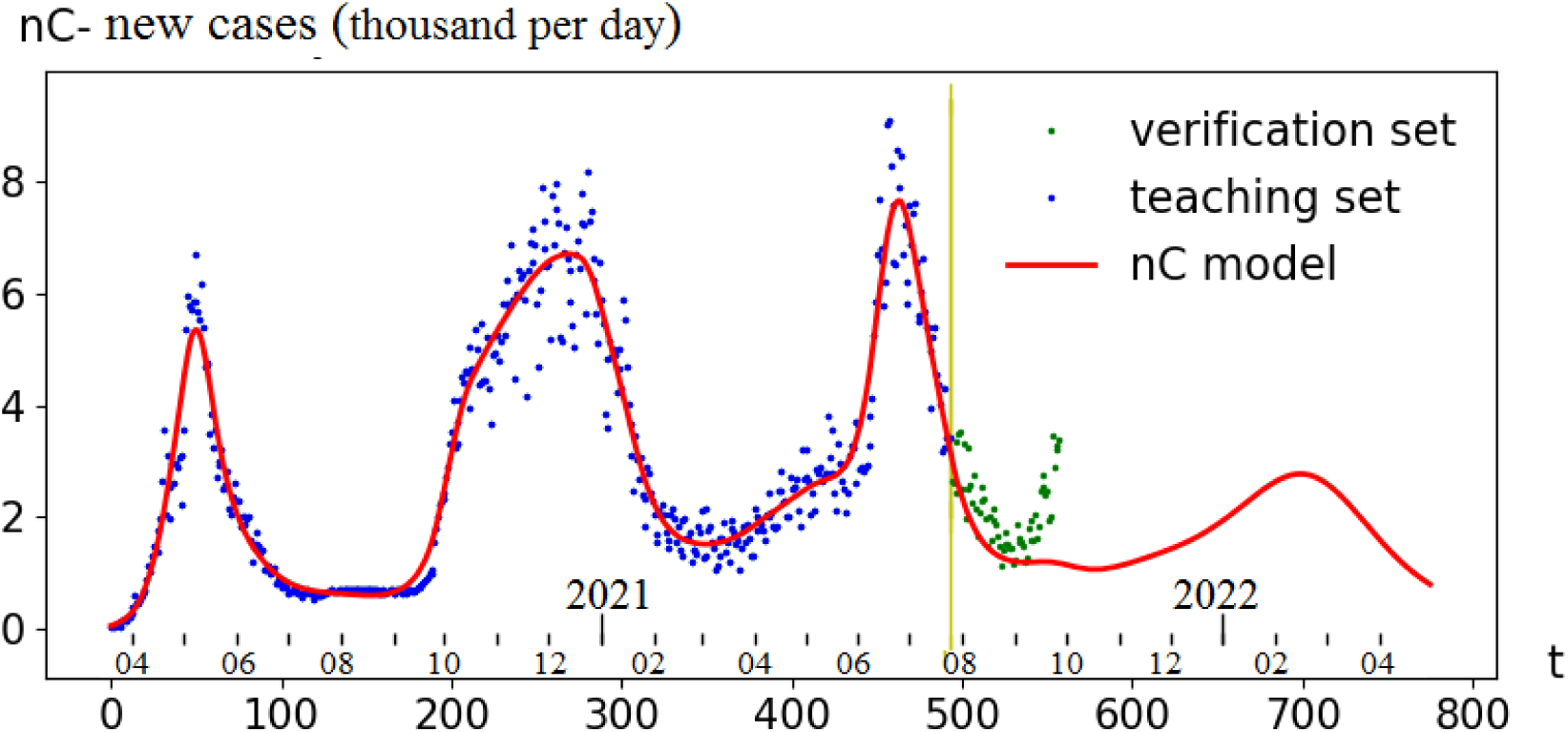
Fourth wave forecast for Moscow from 25.07.2021, and its comparison to real (the green dots) data, up until 27.09.2021

**Fig. 13.**
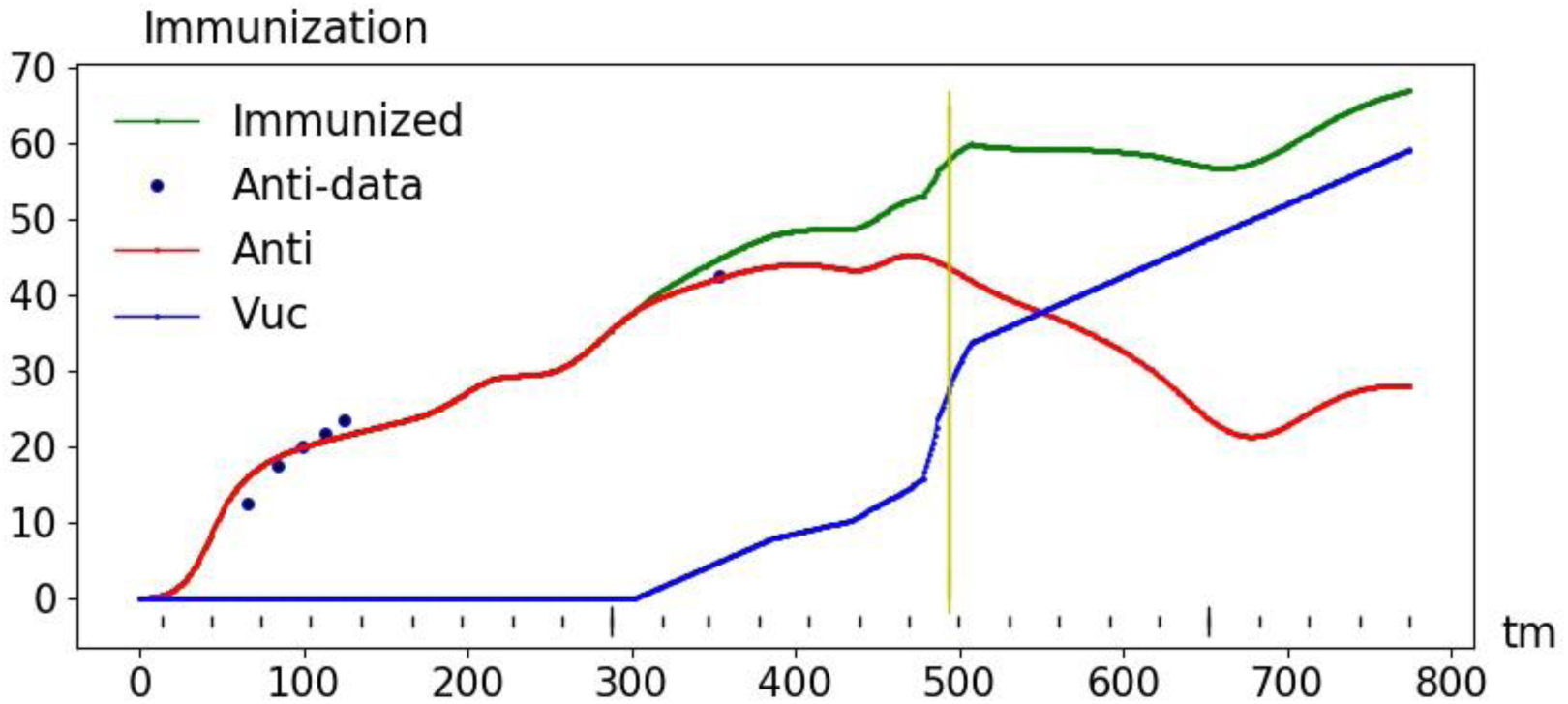
The forecast of immunization of the Moscow population in the fourth wave from 25.07.2021. *Vuc(t)* – preset control, the percentage of vaccinated (corresponds to the rate of vaccination 12 thousand/day). *Anti(t)* – forecast of the percentage of those recovered who retained their immunity by the time *t. Immunized(t)* – forecast of the percentage of the resistant (vaccinated and recovered with immunity)

**Fig. 14.**
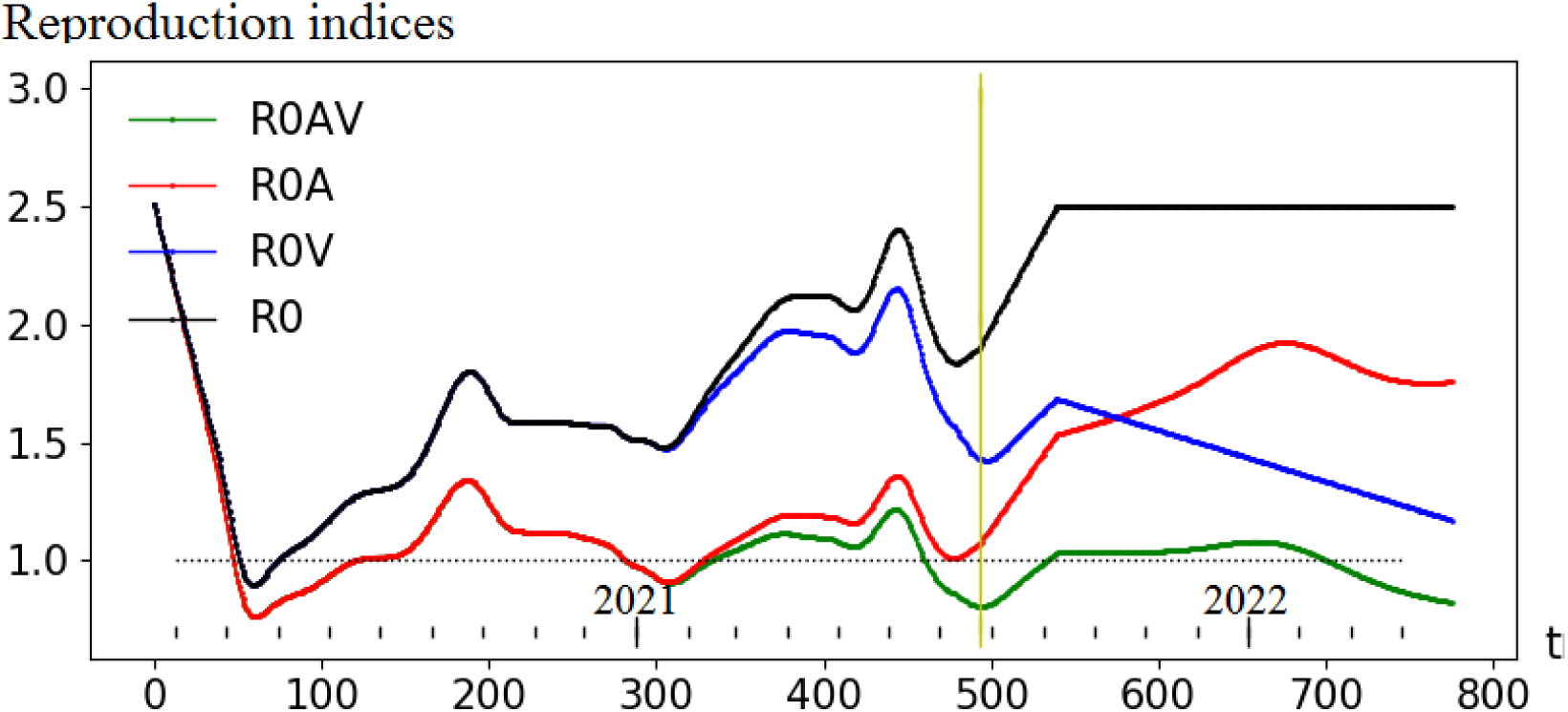
Reproduction indices (as of 25.07.2021): *R0* – reproduction index (identified function and preset control, to the right of the yellow line), *R0A* – reproduction index with the immunity of the recovered factored in, *R0V* – reproduction index with vaccination factored in, *R0AV* – the index of reproduction, with both the immunity of the recovered and vaccination factored in.

**Fig. 15.**
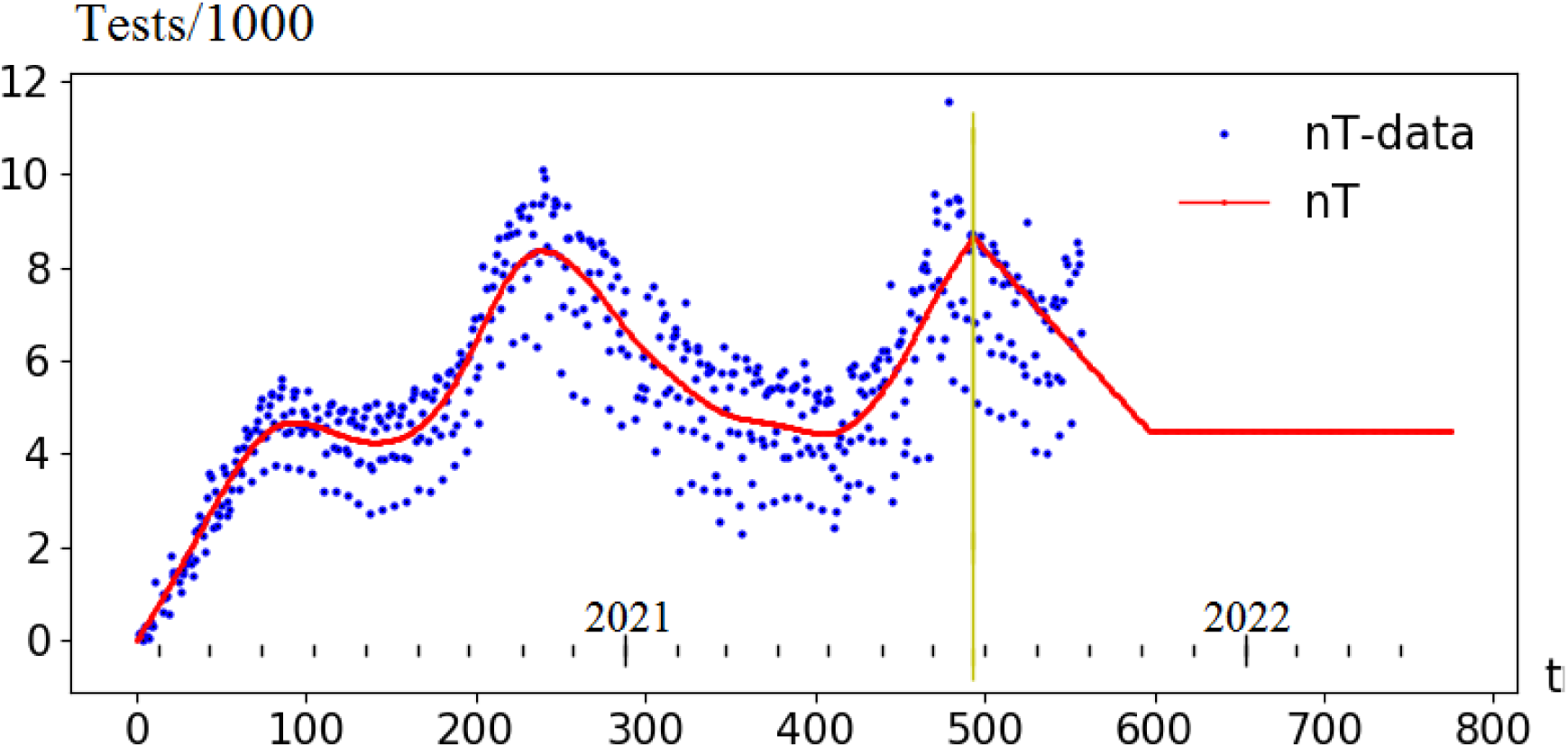
Number of tests per thousand: data fitting and preset scenario for the fourth wave forecast from 25.07.2021 (to the right of the yellow line).

- Vaccination in % of the population of Moscow (in Fig. 13, the *Vuc* graph, shifted by 2 weeks) based on the intensity of 12 thousand/day, the effectiveness is 91.6,
- the reproduction index (R0 in Fig. 14) rises by the beginning of September to a value of 2.5, which corresponds to the absence of restrictions,
- the number of tests per thousand (Fig. 11) by November falls to a moderate value of 4.5
- a small influx of infected from outside of Moscow is expected. The total of 600 per day increases detected infected by about 90 per day.

Fig. 12 shows the forecast for the new cases of infection and real data. For the first month there is decent agreement, but later a noticeable discrepancy appears. It can be explained by a significant increase in the reproductive index due to an increase in infectivity (another strain) and/or by a decrease in immunity of vaccinated over time

## Conclusion

As the COVID-19 pandemic was observed, various effects emerged in the data series studied. Factoring them in made it possible to gradually (i.e. step by step) complicate the model. This process is yet unfinished.

In the result of the monitoring we built a more or less complete (as of August 2021) model that satisfactorily describes the entire annual cycle of the epidemic. The errors that appeared in the forecasts, can mainly be explained by the uncertainty of the scenarios of (future) social administrations required for the calculations.

The current model does not yet factor in:

- Weakening of the immunity of the vaccinated (and subsequent revaccination). By the time the publication was prepared (August 2021), no noticeable manifestation of this process was seen in the accumulated data.
- The migration – the impact of disease rates in the rest of Russia and the world on the disease rates in Moscow. While the dynamics of identified infected was measured in thousands and was determined by internal processes, the migration process, apparently, could be neglected. However, mass vaccination should significantly reduce the number of infected people inside the population (see Fig. 8 and 12) and increase the proportion of the outside flow of the infected. The forecast of the fourth wave from July 25, 2021 with simplified accounting for immigration demonstrated the importance of factoring in this process into.
- Noticeable increase in infectivity due to the virus mutations.

As a result, dynamics for new cases for September 2021 barely fits into the model used, so a new model modification is due. We have to wait until the time series (statistical data) reflect the effects of the loss of immunity from vaccination, the infectivity increase (due to the Delta strain) and the effect of migration becomes noticeable. Apparently, the next significant modification of the model will be needed in October 2021.

*The study was carried out with the financial support of the Russian Foundation for Basic Research within the framework of scientific projects 20-57-82004 and 20-07-00701*

## Data Availability

All data are official statistics. References are given in the text.

## Appendix Mathematical models

### SI_15_R

#### Difference model

According to fig. A1 in this model, the population is divided into 3 main groups: ***S*** – susceptible, ***I*** – infected (detected+undetected) and ***R*** – removed (recovered and isolated). In addition, the infected are divided into 15 subgroups, in accordance with age of infection ***a*** (the time elapsed since the moment of infection)

**Fig. A1.**
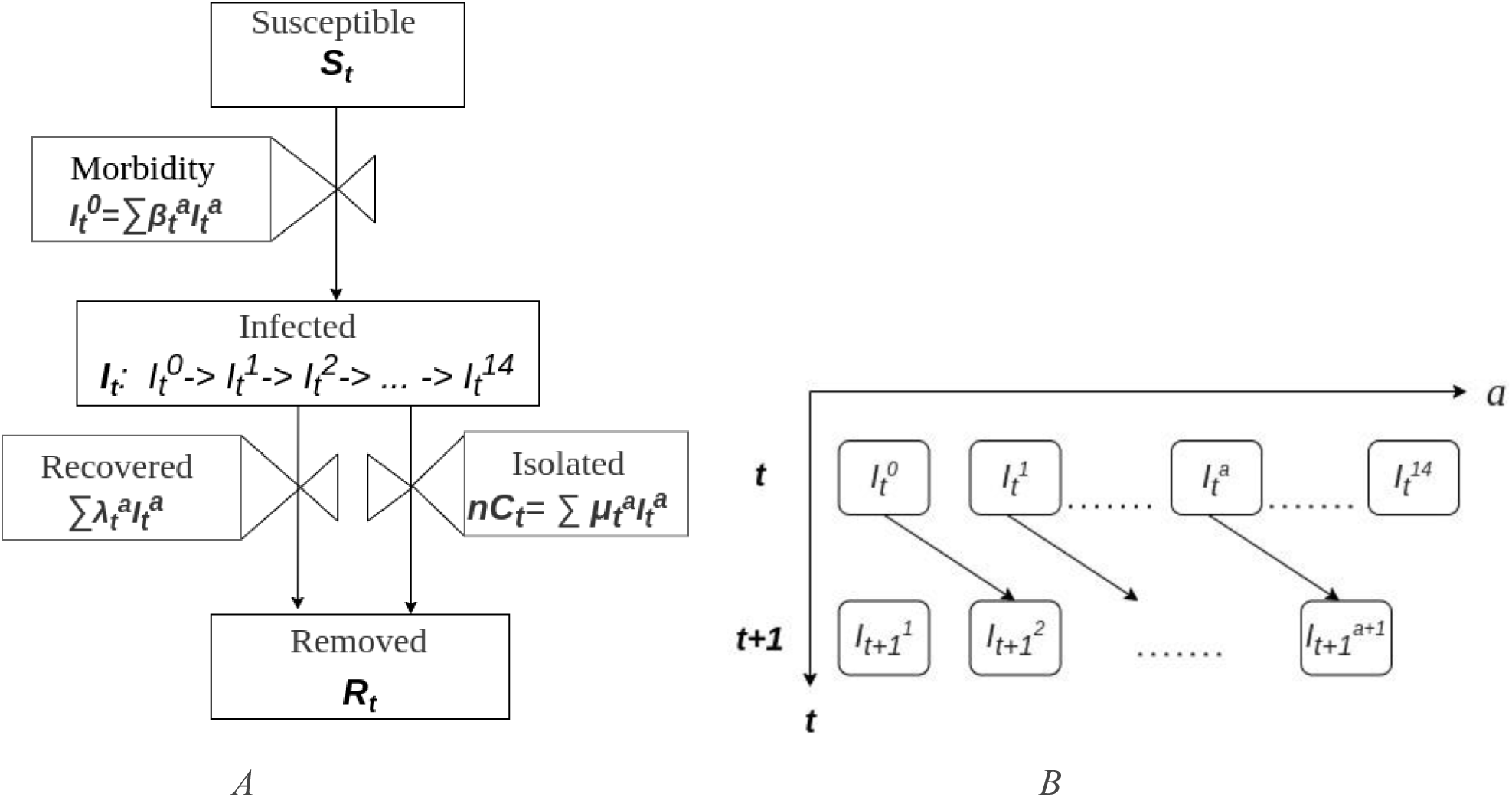
*A*. Model *SI*_*15*_*R: I* (infected) are divided into 15 subgroups. *B*. An illustration of the dynamics of the groups of infected, determined by a shift in the age of infection (***a***).

Formalization of the scheme leads to the difference model:

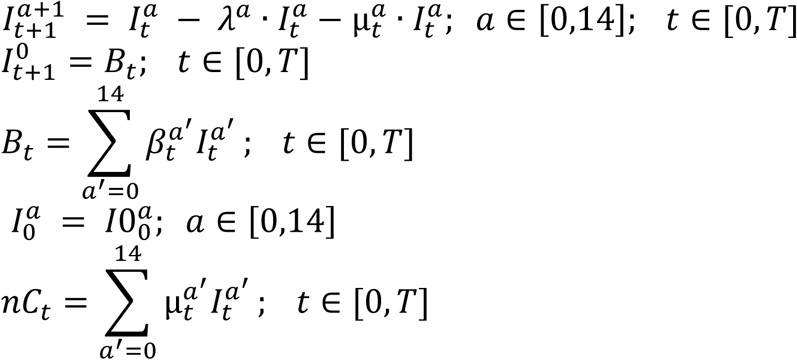

where *a* is age of infection, *t* is the simulation time, 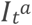 is the number of (undetected) infected at time *t* with the age of infection *a, λ*^*a*^ is the probability of recovering, 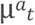 is the probability of detection and subsequent isolation, *B*_*t*_ is the number of new infected, 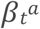 is the infectiousness (contagiousness) of the infected, 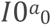 is the distribution of the infected by the age of infection at the start time. *nC*_*t*_ – new (identified) cases of infection.

#### Continuous model

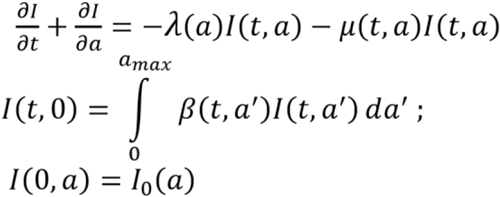

### SI_15_R-nC. New Cases

The main idea of this modification of the *SI*_*15*_*R* model is a multiplicative representation of the functions of detection and infectivity:

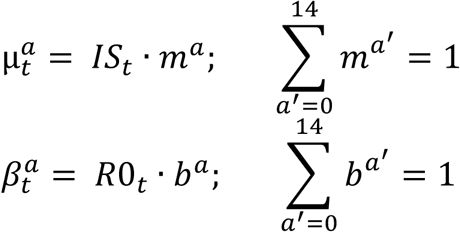

where *IS*_*t*_ is the proportion of the detected and isolated from the total number of infected, *m*^*a*^ is the dependence of detection on the age of infection, *R*0_*t*_ is the current reproduction index or reproductive contact number (how many an infected person infects on average, provided that he is not identified and isolated), *b*^*a*^ is the infectivity as a function of age of infection.

Additionally, rather obvious restrictions on the functions *λ*^*a*^, *m*^*a*^ и *b*^*a*^ are assumed:

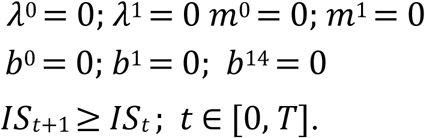

Last inequality (non-decrease in the proportion of detected infected) reflect an increase in society’s efforts to identify and isolate patients (during the first months of the epidemic).

### SI_15_R-nC-nT. New tests

This modification of the model links society’s efforts to identify (and subsequently isolate) the infected to the number of tests performed.

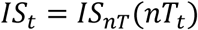

### SI_15_R-nC-nT-Anti. Natural immunity

Taking into account the natural immunity of the recovered (natural herd immunity), number of new infected

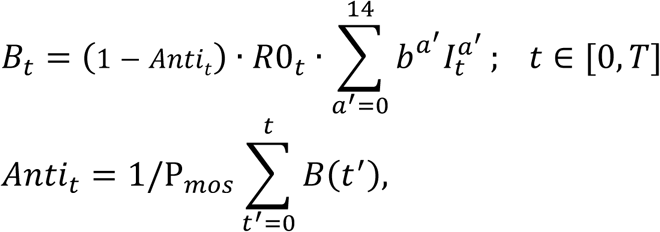

where Anti_t_ – the proportion of antibody carriers at time *t, P*_*mos*_ – population of Moscow.

### SI_15_RS-nC-nT-Anti-Im. Decrease in natural herd immunity

The proportion of antibody carriers

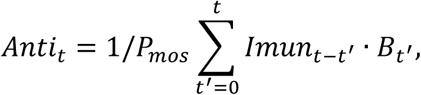

where *Imun*_Δ*t*_ – maintaining immunity as a function of time since the infection (Δ*t*): 1 – complete immunity (the probability of getting sick is 0), 0 – complete lack of immunity.

**Fig. A2.**
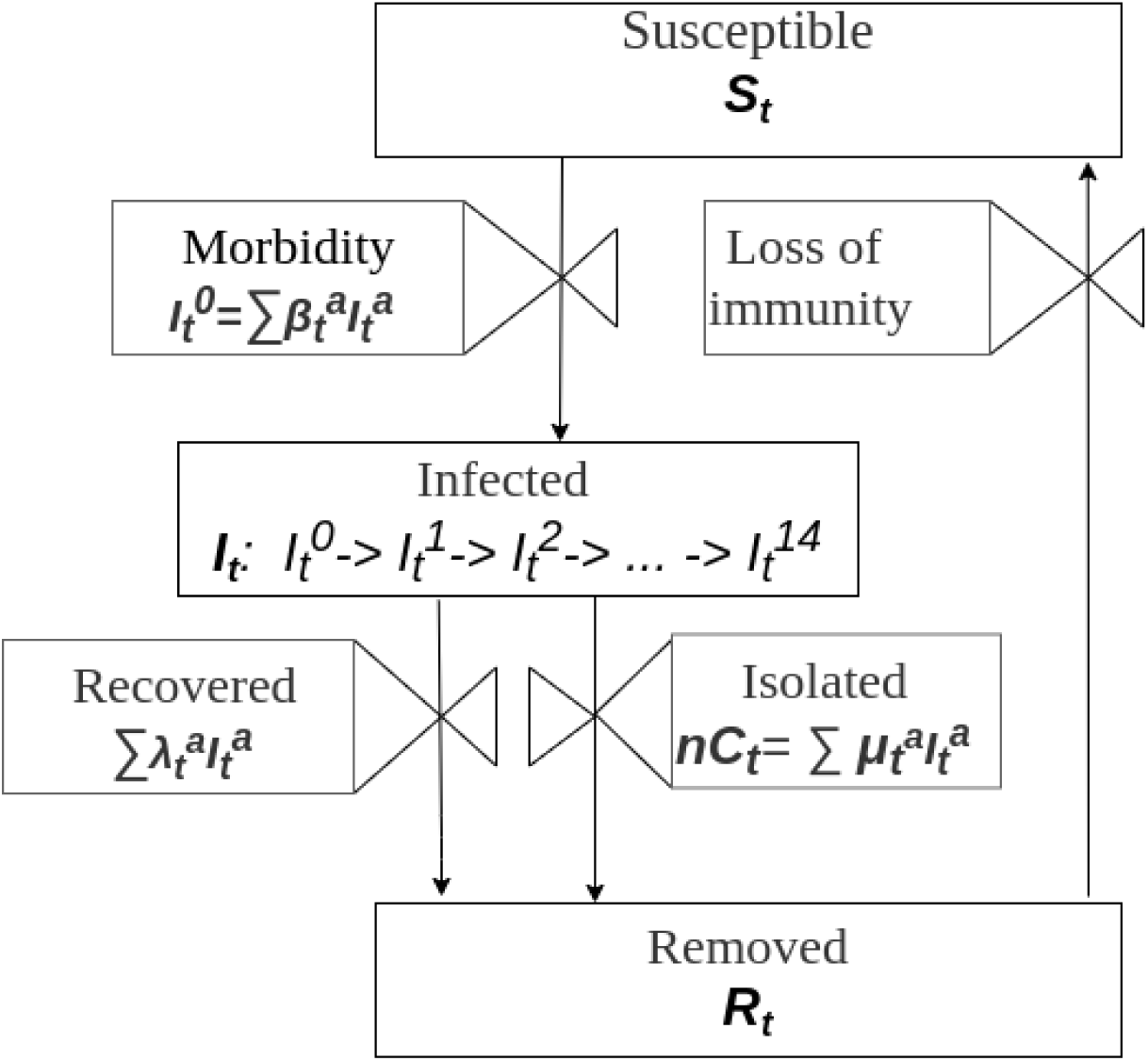
Model ***SI***_***15***_***RS***. The removed (immune) become susceptible again due to loss of immunity.

### SI_15_RS-nC-nT-Anti-Im-Vuc. Vaccination

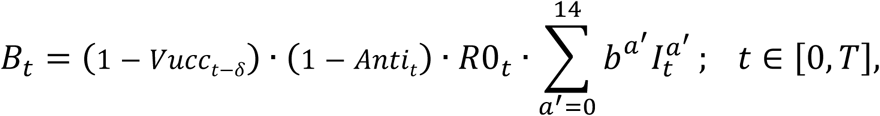

where *Vucc*_*t*_ is the proportion of the vaccinated by the time *t, δ* is the interim between the time of vaccination and the vaccine taking effect (in the calculations, a value of 14 days is used).

